# Systematic review of type 1 diabetes biomarkers reveals regulation in circulating proteins related to complement, lipid metabolism, and immune response

**DOI:** 10.1101/2023.02.21.23286132

**Authors:** Soumyadeep Sarkar, Emily C. Elliott, Hayden R. Henry, Ivo Díaz Ludovico, John T. Melchior, Ashley Frazer-Abel, Bobbie-Jo Webb-Robertson, W. Sean Davidson, V. Michael Holers, Marian J. Rewers, Thomas O. Metz, Ernesto S. Nakayasu

## Abstract

**Aims:** Type 1 diabetes (T1D) results from an autoimmune attack of the pancreatic β cells that progresses to dysglycemia and symptomatic hyperglycemia. Current biomarkers to track this evolution are limited, with development of islet autoantibodies marking the onset of autoimmunity and metabolic tests used to detect dysglycemia. Therefore, additional biomarkers are needed to better track disease initiation and progression. Multiple clinical studies have used proteomics to identify biomarker candidates. However, most of the studies were limited to the initial candidate identification, which needs to be further validated and have assays developed for clinical use. Here we curate these studies to help prioritize biomarker candidates for validation studies and to obtain a broader view of processes regulated during disease development.

**Methods:** This systematic review was registered with Open Science Framework (DOI 10.17605/OSF.IO/N8TSA). Using PRISMA guidelines, we conducted a systematic search of proteomics studies of T1D in the PubMed to identify putative protein biomarkers of the disease. Studies that performed mass spectrometry-based untargeted/targeted proteomic analysis of human serum/plasma of control, pre-seroconversion, post-seroconversion, and/or T1D-diagnosed subjects were included. For unbiased screening, 3 reviewers screened all the articles independently using the pre-determined criteria.

**Results:** A total of 13 studies met our inclusion criteria, resulting in the identification of 251 unique proteins, with 27 (11%) being identified across 3 or more studies. The circulating protein biomarkers were found to be enriched in complement, lipid metabolism, and immune response pathways, all of which are found to be dysregulated in different phases of T1D development. We found a subset of 3 proteins (C3, KNG1 & CFAH), 6 proteins (C3, C4A, APOA4, C4B, A2AP & BTD) and 7 proteins (C3, CLUS, APOA4, C6, A2AP, C1R & CFAI) have consistent regulation between multiple studies in samples from individuals at pre-seroconversion, post-seroconversion and post-diagnosis compared to controls, respectively, making them strong candidates for clinical assay development.

**Conclusions:** Biomarkers analyzed in this systematic review highlight alterations in specific biological processes in T1D, including complement, lipid metabolism, and immune response pathways, and may have potential for further use in the clinic as prognostic or diagnostic assays.

## INTRODUCTION

Our understanding of type 1 diabetes (T1D) pathophysiology has advanced significantly in the last 100 years since discovering insulin as a T1D treatment, but additional biomarkers of its earliest stages could help determine its cause and develop more refined and targeted prevention approaches. The disease begins as an autoimmune insult on pancreatic β cells (seroconversion, marked by the detection of circulating autoantibodies) that progresses to elevated blood glucose and glycated hemoglobin A1c in the body – the current gold standard for T1D diagnosis. Current biomarkers to track this evolution are limited, with the development of autoantibodies to insulin (IAA), glutamic acid decarboxylase (GADA), insulinoma-associated antigen-2 (IA-2A), or zinc transporter 8 (ZnT8A) marking the onset of islet autoimmunity [1, 2]. Proteomics is a powerful tool to identify biomarkers, as it can detect and quantify thousands of proteins. Several proteomics studies have been carried out to identify T1D biomarkers. However, the development of biomarkers is a long process that involves identification of candidates, validation, and clinical assay development [3]. Despite all the efforts of the field, our knowledge is still concentrated in the initial biomarker candidate identification step. A deep analysis of the published reports can recognize reproducible protein expression patterns [4], leading to the identification of most promising candidates.

Here, we performed a systematic review of untargeted and targeted proteomics of serum or plasma from individuals in different stages of T1D development. We report several proteins that were differentially expressed in individuals at various stages of T1D development, and we also interpreted the findings to understand processes regulated in T1D development.

## METHODS

### Study design and search strategy

We conducted this systematic review according to the PRISMA guidelines by searching the PubMed database with the terms “type 1 diabetes” and “proteomics” as of 08 August 2022 [5]. Articles were manually curated with the following expressions:

((Type 1 Diabetes) AND (Proteomics)) NOT ((Review [Publication Type])) OR (Systematic Review [Publication Type])) OR (Meta-analysis [Publication Type]) OR (Commentary [Publication Type])) AND ((Serum) OR (Plasma))

### Eligibility Criteria

Studies comparing the serum/plasma proteome of humans developing or having T1D and that of controls were included in the analysis. Ethnicity, study population size, sex, or disease time point were not included as exclusion criteria to minimize excluding informative biomarkers. We excluded reports of individuals with T1D without matched controls and any studies that failed to report detailed proteomic analyses. Study design (case-control, cohort, or longitudinal) was not an exclusion criterion. We excluded articles without accessible abstracts or full text, articles that were reviews, commentaries, systematic reviews, or meta-analyses.

### Study selection

The systematic review of the literature resulted in 356 initial articles. All the studies that were excluded using the PubMed algorithm (see Eligibility Criteria) were manually verified that they did not meet the inclusion criteria. The remaining articles were manually screened to eliminate studies that did not use human serum/plasma or MS-based proteomic analysis or were related to gestational diabetes and T1D-drug studies. Finally, the studies related to the MS technique without a control group or with missing proteomic data were excluded from the final list after the full text was read. In addition, we added two unpublished manuscripts by our group that met the eligibility criteria. **Figure 1** outlines the study screening and selection process following the PRISMA guidelines.

**Figure 1.**
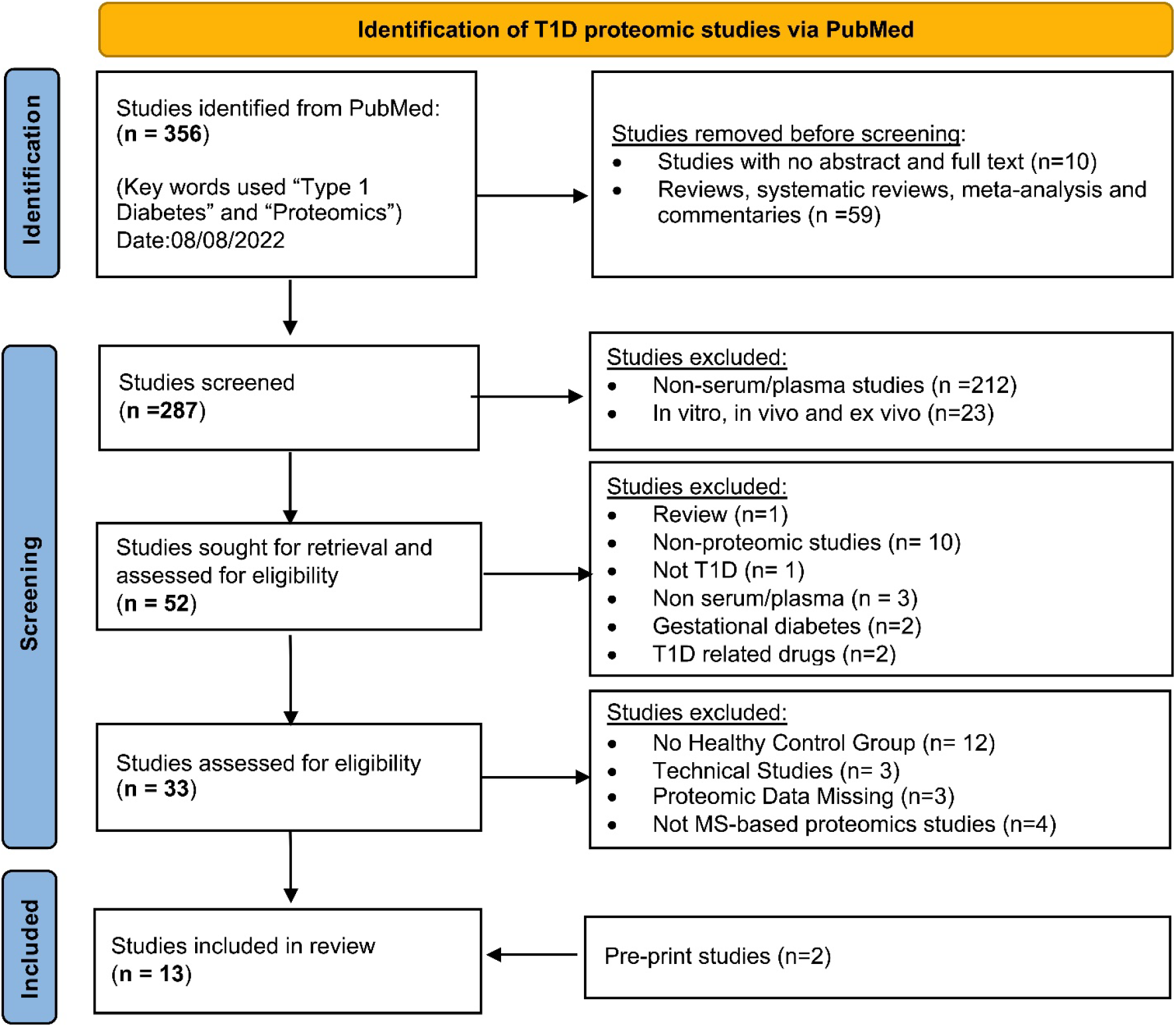
PRISMA flow chart of literature search strategy, screening, and exclusion criteria.

### Data analysis and visualization

The final 13 articles included were screened by three reviewers independently (SS, ECE, HRH) to verify they met the initial inclusion criteria. The additional metadata of sample type, population size, MS-analysis type, method of MS-data analysis, and statistical tests were also considered. All the authors discussed any conflicts and were added to the analysis upon unanimous agreement. Protein data were extracted from the articles manually using Adobe/Microsoft excel and were rearranged using Python and the Python package Pandas [6]. The protein expression data were manually converted to binary “–1” or “1” representing down or up-regulation, respectively, with “0” denoting not reported. Studies were then grouped by the sampling time point (pre-seroconversion, post-seroconversion, and post-diagnosis). Functional-enrichment analysis was performed with Database for Annotation, Visualization, and Integrated Discovery (DAVID) using the default parameters [7]. We used the Kyoto Encyclopedia of Genes and Genomes (KEGG) output for further interpretations. Final data were visualized with Cytoscape (v.3.9.1) and Graphpad prism software.

## RESULTS AND DISCUSSION

### Characteristics and description of eligible studies

The 13 articles described in our systematic review were performed across three disease developmental stages: pre-onset, further divided into (i) pre-seroconversion and (ii) post-seroconversion; and (iii) post-diagnosis. Pre/post seroconversion was defined based on the manifestation of the autoimmune response measured by the appearance of autoantibodies while post-diagnosis was defined by onset of symptomatic hyperglycemia. Details of the studies and their temporal categorization are summarized in **Table 1** and the results are summarized in electronic supplementary material - **ESM 1**.

**Table 1.** Characteristic features of the eligible proteomic studies. A total of 13 studies were identified, and details regarding the various study groups, sampling, and tools for measurement and validation are listed. Red indicates up-regulated proteins, and green indicates down-regulated proteins. Terms used: HLA: human leukocyte antigens, F: Female, M: Male, NHW: non-Hispanic white, DAISY: Diabetes Auto Immunity Study in the Young, TEDDY: The Environmental Determinants of Diabetes in the Young, DASP: Diabetes Antibody Standardization Program, ELISA: enzyme-linked immunoassay, LC-DIA-MS: Liquid chromatography data independent-acquisition-mass spectrometry, LC-MS/MS: Liquid chromatography–tandem mass spectrometry, and LC-SRM-MS: Liquid chromatography-selected reaction monitoring-mass spectrometry

| References | Study group |  | Control group |  | Sample type | Discovery method | Validation method | Number of Proteins Identified by Disease Progression |
| --- | --- | --- | --- | --- | --- | --- | --- | --- |
|  | Clinical definition (Disease condition and sample size) | Demographics (Age and sex, location/ ethnicity) | Clinical definition (Disease condition and sample size) | Demographics (Age and sex, location/ethnicity) |  |  |  |  |
| Pre-seroconversion |  |  |  |  |  |  |  |  |
| Moulder et al, 2015, Diabetes | HLA+, autoantibody+. n=19 | Age (3m to 12y), Finland (DIPP). | HLA+., autoantibody-, controls, n=19 | Age, sex, sample periodicity, and risk group matched | Serum | 2DLC-MS/MS (Untargeted proteomics) | NA | Pre-seroconversion: 3 (1, 2)<br>Post-seroconversion Longitudinal: 55 (30, 25)<br>Post-seroconversion: 10 (5, 5) |
| Frohnert et al., 2020, Diabetes | Family history of T1D, HLA+, autoantibody+. n=20, T1D n=22, | age (AbPos-0.7-26.5y, T1D- 0.7-15y), and sex (AbPos-8F/12M & T1D-10F/12M), Ethnicity (AbPos-15NHW/5BR, T1D-21NHW/1BR), US. (DAISY cohort) | HLA+., autoantibody-. controls n=25 | Matched to HLA genotype, age (0.7-23y), sex (13F/12M), Ethnicity (20NHW/5BR), US. (DAISY cohort) | Serum | LC-SRM-MS, (targeted proteomics) | NA | Pre-seroconversion: 3(1, 2)<br>Post-seroconversion: 2 (1, 1) |
| Webb-Robertson et al, unpublished | HLA+ Pre-seroconversion. n=47, Post-seroconversion. n=131, Pre T1D n=70. | Age (Post-seroconversion~1-23y, pre-T1D ~0-29y), Sex: (Pre-seroconversion-19F/28M, Post-serconversion.- | HLA+. controls n=40 | Matched to HLA genotype, age (~0-14y), and sex (16F/24M), US. (DAISY cohort) | Plasma | LC-SRM-MS, (targeted proteomics) | Exsera | Pre-seroconversion: 8 (0, 8)<br>Post-seroconversion, Post IA: 14 (1, 13) |
|  |  | 63F/68M & pre-T1D<br>-31F/39M), US.<br>(DAISY cohort) |  |  |  |  |  | Post-seroconversion,<br>Pre-T1D: 14 (1, 13) |
| Nakayasu et al., 2022, medRxiv | Untargeted : IA<br>endpoint n=46,<br>T1D endpoint<br>n=46<br>Targeted : IA<br>endpoint<br>n=401, T1D<br>endpoint n=94 | Discovery : T1D :<br>25F/21M<br>IA: 17F/29M<br>Validation :<br>T1D : 43F/51M<br>IA : 179F/222M<br>(TEDDY Cohort) | Untargeted: IA<br>Control n=46,<br>T1D Control<br>n=46<br>Targeted : IA<br>Control<br>n=401, T1D<br>Control n=94 | Matched to clinical<br>center, gender, family<br>history of T1D age, and<br>HLA-DR-DQ genotypes | Plasma | 2DLC-MS/MS<br>(Untargeted<br>proteomics) | LC-SRM-MS<br>(Targeted<br>proteomics) | Pre-seroconversion:<br>C1: 2 (1, 1)<br>Post-seroconversion:<br>C3: 57 (30, 27)<br>Pre-seroconversion,<br>Targeted: 73 (37,<br>36)<br>Post-seroconversion,<br>Targeted: 99 (44,<br>55) |
| Post-seroconversion |  |  |  |  |  |  |  |  |
| Metz et al.<br>(2008) J<br>Proteome<br>Res. | Post-diagnosis<br>n=10 | Age (<30y)<br>(DASP) | HLA-controls, n=10 | Age (<30y)<br>(DASP) | Serum &<br>Plasma | 2DLC-MS/MS<br>(Untargeted<br>proteomics) | NA | Post-Diagnosis: 5 (3,<br>2) |
| Zhi et al,<br>2011, Mol.<br>Cel. Proteom. | Post-diagnosis<br>n=30 | Age (~0 to ~90y),<br>US | autoantibody-<br>controls, n=30 | Age and sex Matched,<br>US | Serum | 2-DE gel-MALDI-TOF MS (Untargeted<br>proteomics) | Luminex and<br>ELISA assays | Post-Diagnosis: 19<br>(11, 8) |
| Chen et al,<br>2012, J<br>Proteomics | Post-diagnosis<br>n=15 | Age and Sex not<br>defined, Taiwan | controls, n=5 | Taiwan | Plasma | LC-MS/MS<br>(Untargeted<br>proteomics) | ELISA and<br>Immuno-blotting | Post-Diagnosis: 36<br>(16, 20) |
| Zhang et al.,<br>2012, J Exp<br>Med | Post-diagnosis<br>n=50 | Age (10-29y), Sex<br>(15F/35M), Mixed<br>ethnicity<br>(DASP) | HLA-., controls,<br>n=100 | Age (18-28y), Sex<br>(51F/49M), Mixed<br>ethnicities<br>(DASP) | Serum &<br>Plasma | LC-MS/MS<br>(Untargeted<br>proteomics) | LC-SRM-MS<br>(Targeted<br>proteomics) | Post-Diagnosis: 24<br>(13, 11)<br>Post Diagnosis<br>Targeted: 24 (5, 19) |
| Manjunatha et al. 2016, Metabolism | T1D-PC n=15 & T1D-GC n=15. | Age (T1D-PC: 33.6 ±12.97y, T1D-GC: 34.5 ± 12.48y), US. | ND-PC n=15 & ND-GC n=15. | Matched for age, sex, and BMI, US | Serum and Plasma | LC-MS/MS (Untargeted proteomics) | NA | Post-Diagnosis: 39 (23, 16) |
| Von Toerne, 2016, Diabetologia | T1D family history, Post-seroconversion, rapid T1D n=15 & slow T1D n=15 | Age (Rapid T1D 0.5-33y, slow T1D 9.5-17.5y), Germany. (BABYDIAB/BABYDIET birth cohorts) | T1D family history, autoantibody-. n=15 | Age and sex matched, Germany (BABYDIAB/BABYDIET birth cohorts) | Serum | LC-MS/MS (Untargeted proteomics) | LC-SRM-MS (Targeted proteomics) | Post-seroconversion: 26 (12, 14) |
| do Nascimento de Oliveira et al, 2018, Diabetes Metab Syndr Obes | Post-diagnosis n=30 | No Familiar history, Age (35.03 ± 8,6), Sex (18F/12F), Brazil. | controls n=30 | No Familiar history, Age (31.5 ± 10.67), Sex (23F/7M), and other clinical criteria matched, Brazil | Serum | LC-MS/MS (Untargeted proteomics) | NA | Post-Diagnosis: 8 (6, 2) |
| Liu et al, 2018, J. Proteomics | HLAPos. Post-seroconversion, T1D n=11. | Age (1-14y), 7 male and 4 female, BR (3) and NHW (8), US (DAISY cohort) | HLA+, autoantibody-. controls n=10 | Age 1-14y, 5 male and 5 female, BR (1) and NHW (9), US (DAISY cohort) | Plasma | LC-MS/MS (Untargeted proteomics) | ELISA | Post-seroconversion: 30 (12, 18) |
| Gourgari et al, 2019, Cadiocasc. Diabetol. | Post-Diagnosis, with high risk of cardiovascular disease, n=26. | 12-21 years old, US (NCT02275091) | controls n=13. | Age, sex, BMI, and clinical lipid measurement matched, US (NCT02275091) | Plasma | LC-DIA-MS (Untargeted proteomics) | NA | Post-Diagnosis: 8 (6, 2) |

#### Pre-onset proteomic profiles

Our literature search identified 6 papers that investigated the temporal protein abundance changes in individuals with T1D. Studies by Moulder et al. [8], Fronhert et al. [9], Nakayasu et al. [10], and Webb-Robertson et al. (unpublished) looked at protein abundance changes at both pre- and post-seroconversion stages. In contrast, von Toerne et al. and Lui et al. examined the protein profile only after seroconversion [11, 12]. Moulder et al., von Toerne et al., Lui et al., and Nakayasu et al. used untargeted proteomics and identified 68, 26, 30, and 59 proteins, respectively, that were significantly different in post-seroconversion vs controls. Nakayasu et al., used targeted proteomics and validated 83 proteins. Webb-Robertson et al. used targeted proteomics, investigating the expression of 24 complement proteins in plasma from pre-seroconversion, and post-seroconversion subjects. In addition, von Toerne et al., and Fronhert et al. used targeted proteomics as a validation method to look at 3 and 5 unique proteins, respectively, whereas Liu et al. used ELISA as its validation step. In conclusion, these studies have identified many biomarker candidates, but with limited validation.

#### Post-diagnosis proteomic profiles

One of the first plasma/serum proteomics studies of individuals with T1D was performed by Metz et al. in 2008 [13]. They identified 5 differentially abundant proteins in recently diagnosed T1D patients compared to controls. Similarly, studies by Zhi et al. and Chen et al. utilized untargeted proteomics and identified 21 and 36 differentially abundant proteins, respectively, in sera from individuals with T1D compared to controls [14, 15]. Zhang et al. and Oliveira et al. performed untargeted proteomics of serum/plasma samples from individuals with T1D and identified 24, and 8 differentially abundant proteins, respectively [16, 17]. Zhang et al. tested the 24 proteins using targeted proteomics in 50 T1D vs. 100 controls, validating 16 proteins with high discriminating power. A subsequent blinded experiment in an independent cohort of 10 individuals with T1D and 10 controls identified the chemokine proplatelet basic factor (PPBP/CXCL7) and C1 inhibitor with 100% sensitivity and specificity to discriminate between the groups. Manjunatha et al. and Gourgari et al. performed untargeted proteomic analysis on high-density lipoproteins (HDL) and found a compositional but not level change of the HDL proteome in T1D individuals with high risk of cardiovascular complications [18, 19]. Overall, these studies showed proteomic changes in plasma profiles after T1D onset, which have the potential to be developed as diagnostic biomarkers.

### Potential biological functions of biomarker candidates

To better understand the biological relevance of these proteins, we performed a functional enrichment analysis using DAVID [7]. The KEGG annotation from DAVID mapped 140 proteins out of 251 to 20 biological pathways (**ESM 2**). Out of these pathways, the complement and coagulation pathway (44 proteins) had the most significant number of proteins mapped to it, followed by COVID-19 (22 proteins), *Staphylococcus aureus* infection (16 proteins), and systemic lupus erythematosus (15 proteins). These pathways were further curated down to 6 major pathways, i.e. complement and coagulation, metabolic protein, inflammatory signaling, cytoskeleton remodeling, extracellular matrix, and antigen presentation, after removing overlapping and redundant proteins, (**Figure 2**). Here, we further discuss these pathways in the context of T1D risk and disease evolution over time.

**Figure 2.**
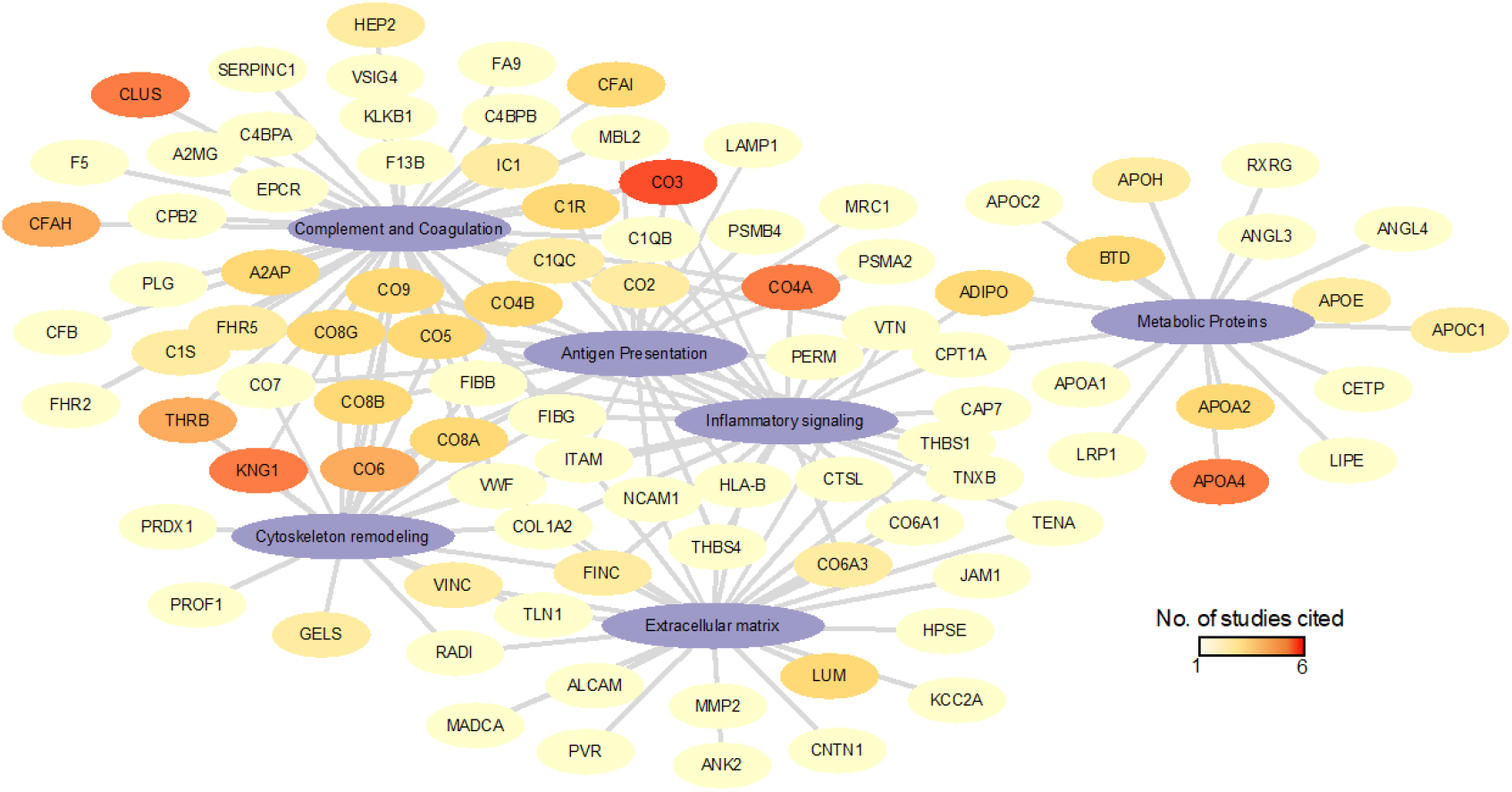
Pathway analysis of the protein biomarkers. This node and string plot representing proteins and their respective consolidated pathways from KEGG. The nodes are colored based on the number of studies that the proteins were shown to be significantly regulated.

**Figure 3.**
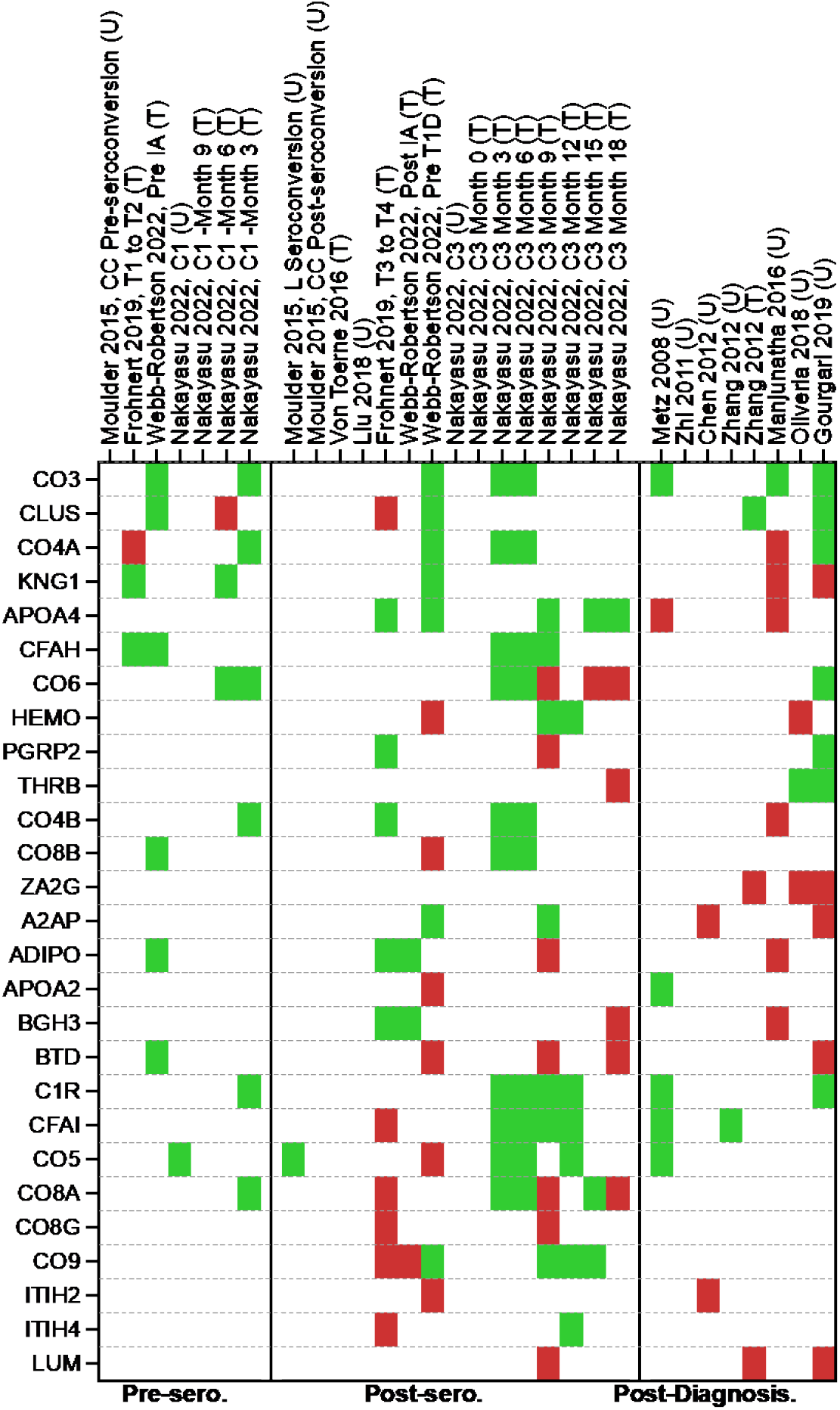
Potential biomarkers list. It is a heatmap of all the protein biomarkers identified by multiple proteomic papers (3 or more times). Upregulated proteins are represented as red and downregulated as green. Proteins that the studies have not reported are represented as blank. “T” denotes the targeted proteomic approach, whereas “U” denotes the untargeted proteomics.

#### Complement system

The complement system is a cascade of proteases making up a humoral extension of the innate immune system. Dysregulation of the complement pathway is linked to chronic and autoimmune diseases. Complement deficiencies are either inherited or acquired. Inherited deficiencies of C1-C4 are strongly associated with bacterial infection and systemic lupus erythematosus, while inherited deficiencies of C5-C9 are associated with bacterial infection and sepsis [20]. Acquired deficiency or factor level changes arise when activation or inflammation-related acute phase responses exist, resulting in either up or downstream exhaustion of some factor [21]. They also interact with adaptive immunity by forming complexes with antibodies, including islet autoantibodies [22]. A cytotoxic effect [23] of such complexes has been controversial, but this concept has gained a renewed interest with recent discovery of β-cell surface autoantibodies [24]. In individuals with T1D, complement components C3 and C4 are highly expressed in the pancreas, including the islets [25, 26]. Studies of pancreata obtained from cadaveric T1D donors have reported C4D immunostaining in blood vessel endothelium and exocrine ducts [25], a change typically associated with activation at that site, and significant upregulation of the complement cascade (C1QA, C1QB, C1QC, C1R, C1S, C3, C4B, C5, C5AR1, C6, C7, C8A, C8B, C8G, and especially C9) [27]. Their tissue compartment localization was extrapolated from transcriptomic data but remains uncertain. This information may be of limited value for our project as the tissue or plasma status of the complement system at the time of death in a person with long-standing T1D may not resemble that at the time of the appearance of islet autoantibodies in an otherwise healthy young child.

Importantly for studies of pre-diagnosis T1D, a functional relationship has been demonstrated between activation of components C3 and C5 and improved β-cell function in mice and humans [28], suggesting direct effects on β cells. The pro-inflammatory cytokines interleukin-1β and interferon-γ increase C3 expression in rodent and human β cells [29]. C3 silencing exacerbates β-cell apoptosis. On the other hand, upregulation of the complement system may improve β-cell autophagic response - a protective homeostatic response to the β-cell stress [29] that is impaired in T1D [30]. Exogenously added C3 protects against cytokine-induced β-cell death via AKT activation and c-Jun N-terminal kinase inhibition. While locally produced C3 is an important survival mechanism in β-cells under a pro-inflammatory assault. However, it is not known if a C3-focused therapy could slow or abort the progression of diabetes in humans. In addition, a variant of C3 gene is associated with T1D [31], which may suggest that the response to such intervention could be genetically modified. In sum, although changes in the complement system are clearly linked to the risk of T1D development and its rapid progression, the direction of the effect and the therapeutic implications are uncertain. Thus, we need to determine whether the system should be activated or modulated, what components of the pathway are most relevant to T1D development, and at what point in the evolution of the disease should a specific change in the pathway be introduced. Regarding complement therapeutics designed to block or modulate activation, there is a range of drugs that are either available or in clinical development. For example, these therapeutics will modulate the C3 and C5 convertases, thus dampening overall activation, or be more specific to target C5, C5a, C3a, or complement receptors for activation fragments (reviewed in [32]). Conversely, activation of the pathways is being explored for the treatment of infectious diseases, cancer, and disorders of metabolism [33].

The 13 papers we examined reported some aspect of the complement system as dysregulated, with 44 out of the 251 biomarker candidates identified from our KEGG analysis (**Figure 4**). C4 was the most identified protein, followed by C3 in this systematic review, and was reported to be primarily downregulated in post-seroconversion and post-diagnosis compared to controls [11, 14, 16, 19]. This was further corroborated by Webb-Robertson et al., where C3 levels were consistently low in pre- and post-seroconverted subjects. This unaltered level of C3 expression throughout the course of the disease was also reported by longitudinal studies conducted by Moulder et al. and Lui et al. These results are consistent with observations of deficiencies in downstream complement components coupled with increased abundance of the MAC inhibitor clusterin pre-seroconversion. However, there are conflicting reports of abundance following seroconversion. Previous *ex-vivo* characterization of the T1D pancreas corroborated our identification of complement dysregulation but instead found an increased abundance of complement markers following diagnosis [25, 27, 34]. Pre-clinical models of C3 knockout or receptor blockage leading to reduction in T1D development and other findings also suggest a role for the complement system in the T1D development [35-37]. While at first, the serum and tissue complement abundances appear to be at odds with one another, the low complement levels pre-seroconversion may be due to consumption through C3, C4, and MAC depositing in the pancreas throughout T1D development. Low C3 and C4 have been seen in both COVID-19 and lipodystrophy cases, and low C3 with high C5-C9 are common in glomerulopathy [38]. Evidence of lifelong complement deposits in the pancreas matching the parallel findings of lifelong low C3 levels provides further evidence of complement deposition in the pancreas as the driving force behind low blood levels of complement and, therefore, progressive loss of β cells due to increased immune response.

**Figure 4.**
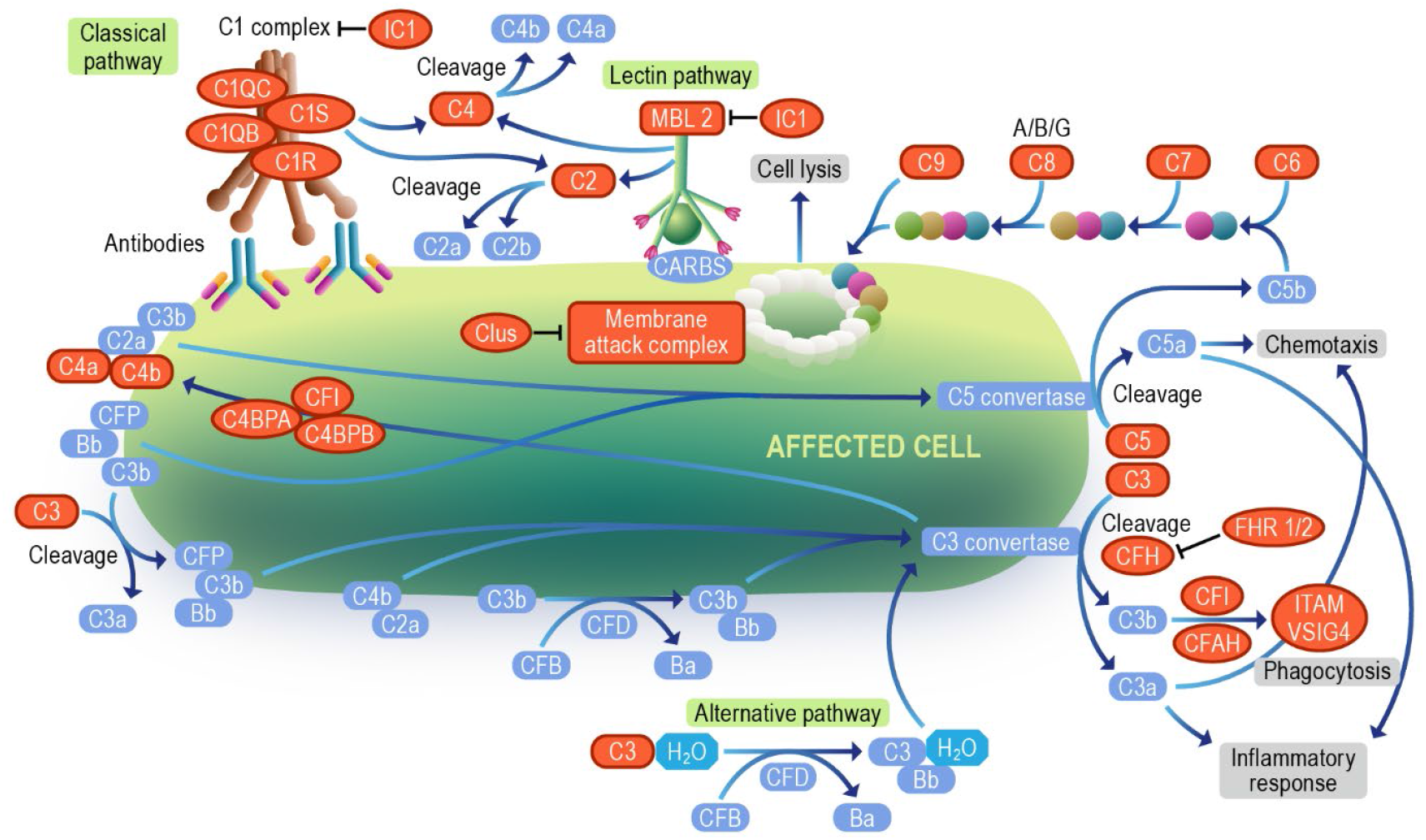
Complement cascade. The diagram represents all the complement pathway proteins denoted by orange color identified in our systematic review. The Image was modified from “Complement cascade pathway” on the Reactome website (https://reactome.org/PathwayBrowser/#/R-HSA-166658 with StableID: R-HSA-166658).

In the context of T1D, the complement system may play a part in modulating adaptive immune response against islets, with HLA class II genes being associated with T1D risk [39]. Activation of the complement cascade and deposition of complement factors into the pancreas have been reported during insulitis [22, 27]. In late-stage diabetes, elevated complement levels in serum are linked to diabetic nephropathy [40]. Overall, the disruption of complement pathway seems to be a characteristic trait of T1D, however, further studies are warranted to help navigate the path to consistent biomarker or drug development.

#### Immune pathways

A recurrent theme among the T1D biomarker candidates is the enrichment of proteins related to antigen processing and presentation. For instance, the complement pathway can opsonize pathogens and dead cells to be presented to the antigen-presenting cells. Other proteins related to antigen opsonization are also regulated in T1D, such as antibodies and opsonization/scavenger receptors. Once the antigen is opsonized, it is phagocyted into phagosomes. In fact, β cells infected with coxsackievirus are efficiently phagocyted by dendritic cells, making coxsackievirus infection a potential trigger of the islet autoimmune response [41, 42]. The phagocytic process requires an extensive cell cytoskeleton remodeling [43], which was another pathway enriched in our analysis. The phagosome can be next fused to lysosomes to initially process the antigens, which are further processed in the proteasome and loaded into HLA for presenting to T cells [44]. HLA alleles represent the main risk factor of T1D development, further supporting that this pathway is involved in the autoimmune response [44].

Another process that occurs in parallel is the cytokine and chemokine signaling [45]. Among the biomarker candidates, the chemokine PPBP/CXCL7 has been identified along with 3 other immunoregulatory molecules (**Figure 5**). Despite all the cytokines/chemokines regulated in T1D, little is known about their mechanistic roles in disease development. Cytokine/chemokine and even phagocytosis can trigger signaling cascades in the cells that further regulate these processes but also leads to the expression of other effector molecules. Among signaling proteins, 4 kinases, phosphatases, and phospho-binding proteins have been described (**Figure 5**). In addition, the transcription factor TNIP1 has also been shown to be regulated in T1D (**Figure 5**). Regarding the effector molecules, oxidative stress proteins myeloperoxidase, glutathione peroxidase 3, peroxiredoxin-1, and sulfhydryl oxidase 1, were also shown to be regulated in T1D. Oxidative stress has been shown to induce β-cell dysfunction and death and has also been proposed as a potential therapeutic target [46].

**Figure 5.**
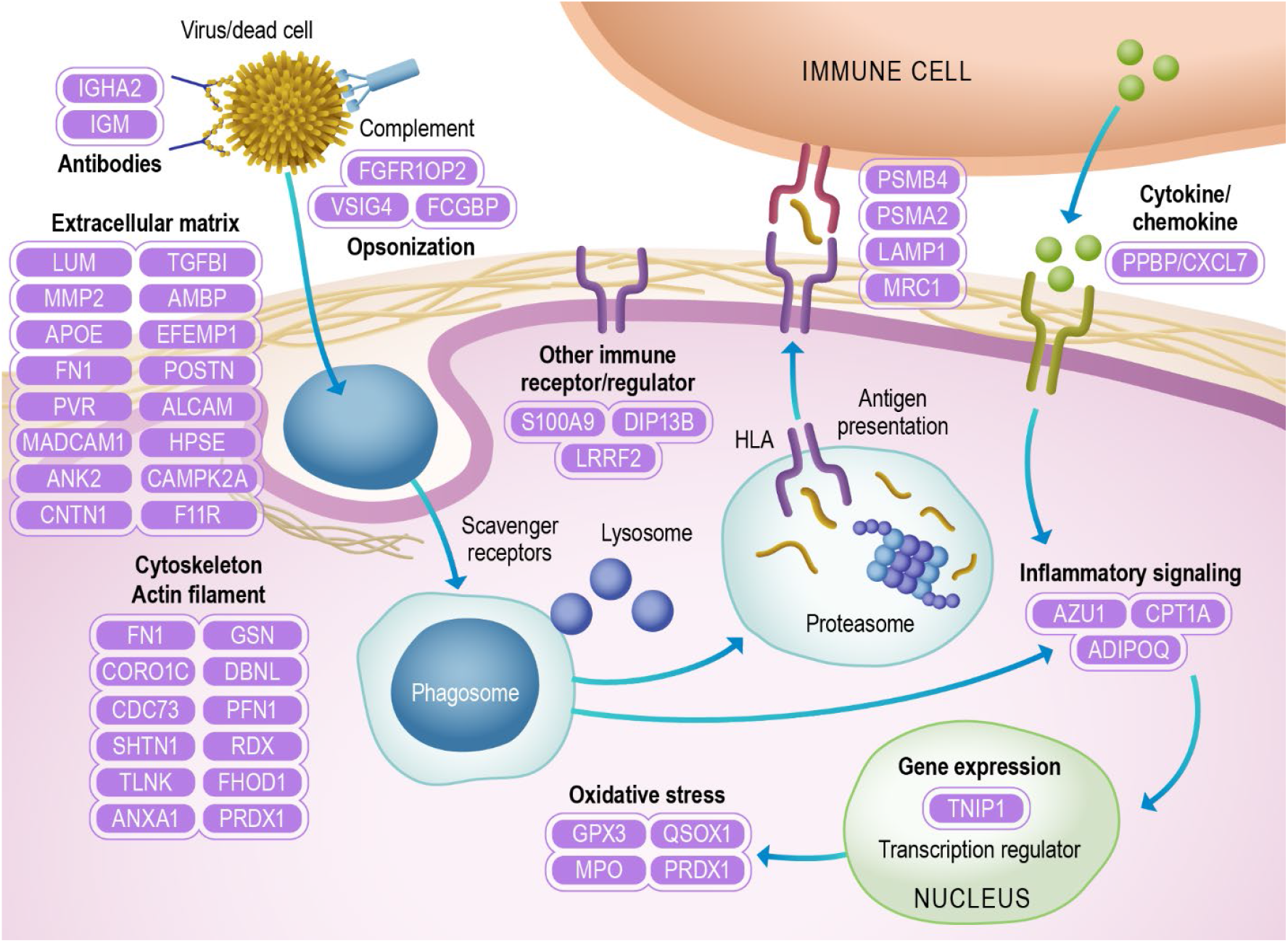
Immune pathways. The diagram represents proteins identified as the extracellular matrix, cytoskeleton/actin filament, oxidative stress, gene expression, inflammatory signaling, antigen presentation, cytokine/chemokine, opsonization, antibodies, and other immune receptors/regulators.

A common feature in the plasma of individuals with T1D is the regulation of extracellular matrix proteins, of which 27 have been described to be regulated (**Figure 5**). The extracellular matrix is an integral part of the immune response regulated by cytokines and chemokines [47, 48]. For instance, the extracellular matrix peri-islet basement membrane serves as a barrier, protecting islets from immune cell infiltration in insulitis in mouse models of T1D [49]. In addition, preservation of the extracellular matrix by administration of dextran sulfate, a mimic molecule of the extracellular matrix proteoglycans, has been shown to protect β cells and to be a potential treatment for T1D in mice [50].

#### Plasma lipoproteins

The DAVID analysis showed ∼5% of the reported proteins to be key players in lipid metabolism. Most circulating plasma lipids are packaged into lipoproteins which are traditionally classified into four common subfractions based on particle density: chylomicrons (CM), very low-density (VLDL), low-density (LDL), and high-density lipoproteins (HDL). Structurally dynamic apolipoprotein scaffolds reside at the water-lipid interface of all subclasses where they modulate particle interactions with plasma enzymes, cofactors, and cell surface receptors that continuously remodel the lipoproteins throughout their lifespan. Though traditionally defined based on their “cholesterol” content, proteomics studies over the last decade have revealed significant compositional heterogeneity exists within the lipoprotein subfractions which contain upwards of 273 different proteins [51] with the HDL subfraction accounting for >250 of these proteins. Thus, lipoproteins are thought to consist of a variety of compositionally distinct subspecies which have now been shown to modulate a diverse array of metabolic pathways [52, 53].

Though DAVID analysis implicated “cholesterol metabolism,” and by proxy lipoproteins, the analysis fails to capture the full lipoproteome due to the recency of lipoprotein molecular profiling studies in the literature. When compared to a lipoprotein-specific database [51], we found nearly half (N = 112) of the protein biomarkers identified in individuals with T1D are associated with lipoproteins (**Figure 6A, ESM3**), and 66 were more specifically associated to HDL (**ESM4**). Approximately 85% of these changes were unique to post-seroconversion and post-diagnosis indicating the most profound changes in lipoprotein metabolism occurs later in the disease process (**Figure 6B)**. A total of 17 members were altered pre-seroconversion with most overlapping with previously discussed immune response and complement cascade (**Figure 6C**). Outside of the immune and complement proteins, we noted a few well-studied APOCs and clusterin were altered pre-seroconversion (**Figure 6D**), hinting some changes occur in traditional lipoprotein metabolism pathways prior to onset of dysglycemia or hyperglycemia.

**Figure 6.**
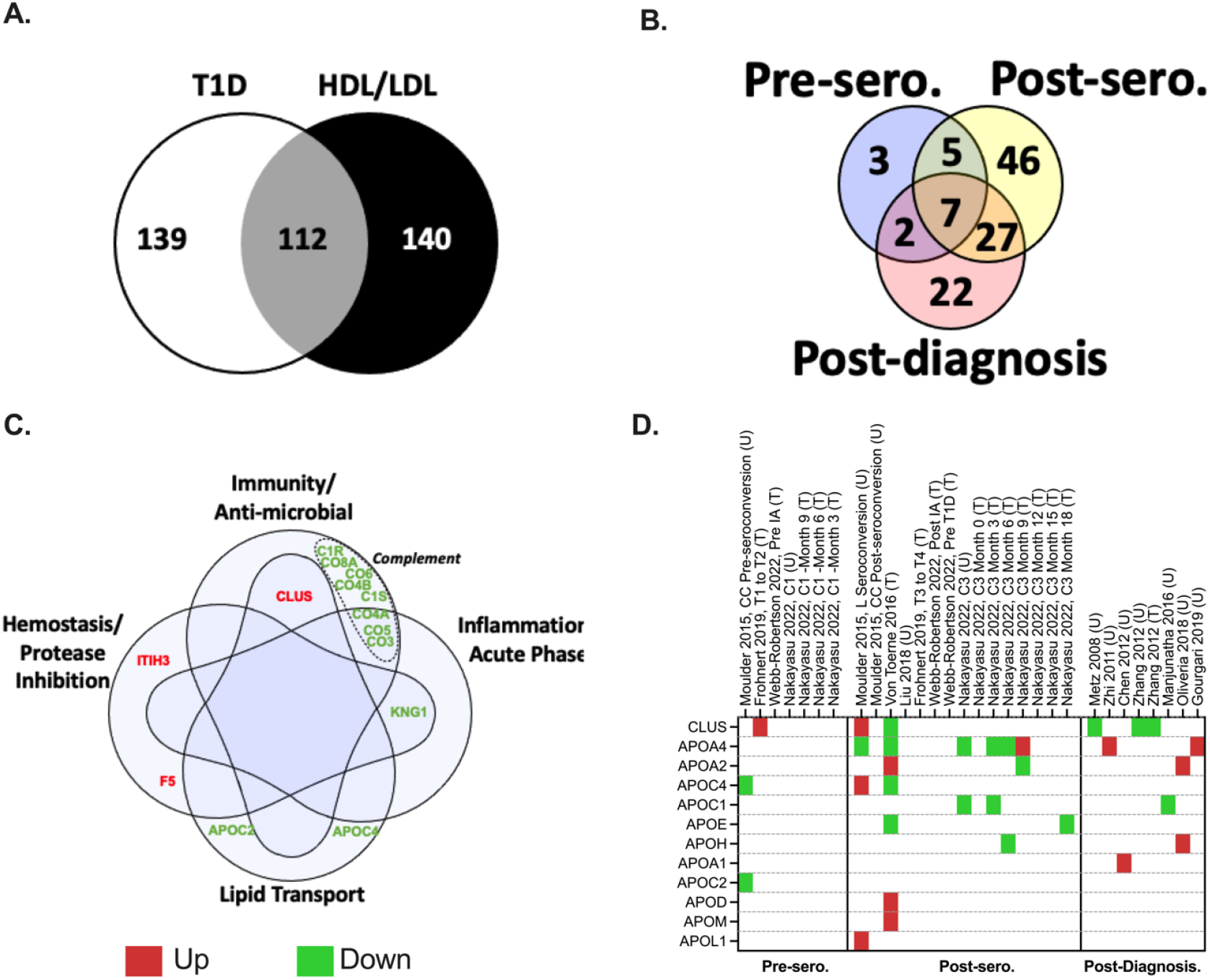
Overlap of T1D-relevant proteins with HDL/LDL lipoproteome. A. Venn diagram showing common protein hits (grey) between the HDL/LDL proteome [51] (black) and those reported in our systematic review (white). **B**. Venn diagram detailing the T1D stage as pre-seroconversion (pre-sero), post-seroconversion (post-sero) and post-diagnosis of the 112 HDL/LDL/T1D shared proteins of figure A. **C**. Gene Ontology of HDL lipoprotein-known functions of pre-seroconversion reported proteins. **D**. Heatmap of apolipoproteins with altered levels reported in at least one stage of T1D development. Panel C was modified from a published figure by Davidson et al. (copyright permission was obtained from the publisher, license number: 5433910818199) [51].

Most of the lipoproteome members altered post-seroconversion and post-diagnosis have documented roles in triacylglycerol metabolism. Perhaps the most robust of these observations was the associated with changes in plasma APOA4 altered in 8 studies post-seroconversion and post-T1D diagnosis [8, 11, 14, 18, 19]. APOA4 is well-documented to modulate the triacylglycerol packaging in the triacylglycerol-rich lipoproteins (CMs and VLDL). [54]. Additionally, APOA4 is reported to play key roles in satiety, gastric function, and glucose homeostasis [55, 56], all of which have been reported altered in individuals with T1D [56-60]. Two post-seroconversion and post-diagnosis studies reported increased plasma APOA2; a well-known HDL scaffold protein. While HDL which has little triacylglycerol, APOA2 has been shown to be implicated in triacylglycerol metabolism [61] though the mechanism is still poorly understood. Several studies report changes in the APOCs and APOE [8, 11, 18] which are also thought to modulate triacylglycerol lipolysis on VLDLs [62, 63]. These observations are in-line with elevated triacylglycerol levels and inhibition of lipoprotein lipase associated with the innate immune response [64].

Most of the biomarker studies were performed on whole plasma. As most apolipoproteins are exchangeable, more detailed lipoprotein speciation studies are required to determine the subclass on which these particles are located and how they are modulating particle function in the context of T1D. Though two studies attempted to speciate lipoproteins from individuals with T1D, both limited their analysis to HDLs (21, 22) thus missing most of the changes involved with the triacylglycerol-rich particles. Future studies that examine the temporal changes in the lipoproteome across all subclasses will better inform on the role of these pleiotropic particles and how they cross-communicate with the aforementioned complement system and immune pathways in T1D.

### Overview of the most promising biomarker candidates for clinical use

For clinical use, biomarkers need to be highly sensitive and specific. As T1D is a chronic disease in which each stage can take months to years, biomarkers with more stable changes in abundance are preferable to proteins with transient regulation. In addition, they need to be reproducible. Therefore, proteins with consistent abundance profiles across different studies would make them stronger candidates to be developed into clinical assays. Our systematic review identified 251 proteins, 66 (26%) were found across multiple papers, and 27 (11%) were reported three or more times (**Figure 3**). The differential expression patterns were more limited in the pre-seroconversion than post-seroconversion, with most differentially expressed proteins in the post-diagnosis group. Upon evaluating the directional expression (i.e., up and down) of the biomarker proteins across all three temporal groups (pre-seroconversion, post-seroconversion, and post-diagnosis), complement proteins C3, C6, and C1R were found to be consistently downregulated in cases compared to HC. Eight proteins (C4B, APOA4, C2, BGH3, ADIPO, A2AP, IBP2, and GPX3) identified as low expressed in pre/post-seroconversion individuals were up in post-diagnosed individuals, whereas 5 proteins (SAMP, CBPN, ITIH2, FHR5, & PA2G4) which were up in post-seroconversion stage, were down in post-diagnosed individuals. A subset of proteins was only identified in the post-seroconversion group (C5, C1QC & CFAI) or post-diagnosis group (THRB, ZA2G, HPT, CBG, & LUM) by 2 or more articles. Therefore, these biomarkers are strong candidates for further validation studies and the development of clinical assays.

### Conclusions and perspectives

Our systematic analysis found 251 candidate protein biomarkers of T1D, of which 66 (26%) were observed in multiple studies. This helps to prioritize for the validation step of biomarker development. Despite some of the biomarkers being consistently regulated across different studies, they still need to go through an extensive validation process before moving to the clinical assay development. Ideally, the biomarker candidates should be cross-validated in independent cohorts of samples and tested for sensitivity and specificity. In general, fewer signatures were identified prior to the onset of the disease. This can be partly because T1D has an almost silent developmental phase, and it is not expected that significant biochemical changes would be observed in the blood of these individuals. Alternatively, the pre-seroconversion phase may be convoluted by multiple factors and trajectories that lead to autoimmunity and hinder our ability to identify a consistent signature. In this context, machine learning can be an excellent approach to identifying multivariate panels of proteins to serve as biomarkers of T1D development. This approach has been used to combine metabolic, genetic, and autoimmune signatures to predict the onset of disease and can be easily adapted to test peptide/protein panels [9, 10, 65, 66]. Another concept that can further improve biomarkers’ robustness is using ratio between protein abundance changes rather than profiling individual proteins. Ratios between oppositely regulated proteins would have much bigger differences compared to them individually, providing higher discriminatory power between cases and controls. After selecting candidates, clinical-grade assays must be developed and tested for robustness, specificity, and sensitivity in the clinical setting. In addition to biomarkers, our systematic review of proteomics studies provided insights into the pathways regulated in T1D, such as complement system, plasma lipoproteins, and immune response [65, 66]. Our systematic review also opens opportunities to study the functions of the biomarker candidates in T1D development and pathology. For instance, our group has found that PPBP/CXCL7 can reduce pro-inflammatory cytokine-mediated apoptosis in macrophage cell cultures while it enhances it in cultured β cells [67]. This may have a role in T1D development by potentiating macrophages and killing β cells in insulitis. Overall, this systematic review provides insights into processes regulated in T1D development and highlights some of the best candidates for developing clinical assays.

## Data Availability

The study design was registered at Open Science Framework.

https://osf.io/n8tsa/

## SUPPLEMENTARY MATERIAL

Supplementary material is electronically available with this manuscript and on Open Science Framework (OSF) website https://osf.io/n8tsa/

## ACKNOWLEDGMENTS

The authors thank Mr. Nathan Johnson of PNNL for his help drafting figures.

## FUNDING

The Leona M. and Harry B. Helmsley Charitable Trust (to M.R.). National Institutes of Health, National Institute of Diabetes and Digestive and Kidney Diseases grants U01 DK127786 (to T.O.M. and B-J.W-R.) and U01 DK127505 (to E.S.N).

## CONFLICTS OF INTEREST

The authors declare no financial conflicts of interest.

## Notes

### Competing Interest Statement

The authors have declared no competing interest.

